# The Transition from adolescence to adulthood mental health services for young people with ADHD in Italy

**DOI:** 10.1101/2024.05.31.24308270

**Authors:** E. Roberti, A. Clavenna, E. Basso, C. Bravaccio, M. P. Riccio, M. Pincherle, M. Duca, C. Giordani, F. Scarpellini, R. Campi, M. Giardino, M. Zanetti, V. Tessarollo, I. Costantino, TransiDEA Group, M. Bonati

## Abstract

**Aims:** Ensuring a successful transition to Adult Mental Health Services (AMHS) is fundamental for ADHD patients to prevent adverse scenarios in adults (e.g., psychiatric disorders, substance or alcohol abuse). Yet, most European nations do not have appropriate transition guidelines. This study aims to enquire about the current transition paths in Italy and the perceived experiences of the patients and their clinicians.

**Methods:** The present qualitative, observational study collected 36 interviews with young adults with ADHD who turned 18 between 2017 and 2021. Simultaneously, two questionnaires were filled in by the clinicians (both from pediatric and adult mental health services) who were involved in their transition paths. These tools collected information about the transition process, the services that cared for the young adults, and well-being indicators such as impairment in daily life, employment status, and the presence of sentinel events (e.g., critical stage accesses to the emergency room or hospitalizations). Successful and failed referrals were analyzed.

**Results:** A referral to an AMHS was attempted for sixteen young adults (8 before age 18 and 8 when turning 18), and 8 patients (22.2% overall) were successfully taken into the care of the AMHS. Twenty patients were not referred since it was deemed unnecessary (*N*=6) or because of the lack of specialized services or compliance (*N*=14). At the time of the interview, only nine participants were still under AMHS care. Of eleven individuals with a high need for care (identified by the level of impairment, support needs, or sentinel events), five were not followed by a mental health professional at the time of the interview.

**Conclusions:** For the majority of ADHD young adults, a transition path was never started or completed. While this is partly due to mild levels of impairment, in many cases it was difficult to find a service that could care for the adult patient. Only 1 out of 4 young adults are successfully transferred to AMHS care. Creating or improving evidence-based transition guidelines should be a priority of the public health system to ensure healthcare for as many patients as possible. The results of this study will converge towards the need for recommendations for the transition of services from adolescence to adulthood for young people with ADHD for Italian clinical practice.

## Introduction

Attention Deficit/Hyperactivity Disorder (ADHD) is a neurodevelopmental disorder with a prevalence of around 5% in childhood and adolescence (Seixas, Weiss and Müller, 2012; Roughan and Stafford, 2019). About 7% of the patients continue to have impairing symptoms in adulthood (Song *et al*., 2021). Moreover, ADHD in adults has a high comorbidity with psychiatric disorders, substance or alcohol abuse, antisocial personality, or even criminal activities (Di Lorenzo *et al*., 2021). To prevent the onset of adverse scenarios, it is fundamental not to lose patients when they stop being cared for by Child and Adolescent Mental Health Services (CAMHS). Instead, too often, the apparent lack of severe symptoms hinders a successful transition to Adult Mental Health Services (AMHS) (McCarthy *et al*., 2009; Young, Murphy and Coghill, 2011).

Several factors contribute to this discontinuity of care in adolescence: lack of organization, scarce resources and collaboration between services, stigma, concerns about peers judgement, and low family compliance (Anderson, Newlove-Delgado and Ford, 2022; Roberti *et al*., 2023b). The result is feeling left alone (Driver *et al*., 2022) and a later worsening of symptoms with urgent service re-entries (Anderson, Newlove-Delgado and Ford, 2022).

While some guidelines exist at an international level (e.g., the NICE Transition Guidelines and the Ready Steady Go program in the U.K., The Six Core Elements in the U.S.A.) (National Institute for Health & Care Excellence, 2016; White *et al*., 2018; Meyers and Irwin, 2023), their feasibility at a practical level is questioned (Signorini *et al*., 2017; Eke, Janssens and Ford, 2019; Eke *et al*., 2020). Moreover, not all countries possess such regulations, as is the case for most European nations. The improvement of transition guidelines should be a priority of the healthcare system. This need is particularly evident and urgent in Italy: in a country where the estimated ADHD prevalence ranges from 1.1 to 3.1% among youth aged 5 to 17 years (Reale and Bonati, 2018), no shared transition practices by CAMHS and AMHS currently exist. This observation emerged within the first phase of the *Transition care between adolescent and adult services for young people with chronic health needs in Italy* project (TransiDEA – Transition in Diabetes, Epilepsy, and ADHD patients), designed to assess the current state of transitioning practices, the experience of patients, their families and the clinicians of both CAMHS and AMHS (Roberti *et al*., 2023b). Indeed, amongst forty-two services that participated in the 2022 Survey (Phase 1), only twenty-one declared having a transition protocol, and even fewer (six in total) provided a copy. A fragmented picture emerged from the analysis of such protocols (Roberti *et al*., 2023a). The goal of the present study (Phase 2) was to enquire about the paths and perceived experiences of the patients and their CAMHS and AMHS (in case of a successful transition) clinicians. Putting together the information collected in phases 1 and 2 will lead to the definition of a consensus document to be adopted in Italian services (Phase 3).

## Methods

The present observational qualitative study was conducted between January and July 2023 as a part of the TransiDEA project - ADHD branch. The goal was to collect at least 30 semi-structured interviews (10 from the north, 10 from the center, and 10 from the south of Italy) of young people who turned 18 within the past 3-6 years (approximately 2017 to 2021). The interviews aimed to collect information about patients when they turned 18, in the year before the interview, and in the intermediate period (i.e., the time between turning 18 and the year preceding the interview) (Figure 1). In parallel, two questionnaires were developed for the CAMHS and AMHS clinicians involved in the transition paths. These tools were designed based on Phase 1’s results and discussed with a small group of clinicians, and adjustments based on their feedback were made. Specifically, the content of the interview enquired about:

a. ADHD type, comorbidities treatments, and impairment areas in daily life;
b. Services that cared for the young patient before turning 18, in the year before the interview, and in the intermediate period;
c. The severity of symptoms, treatments, and occupation at the time of the interview, as well as the presence of sentinel events (critical stage accesses to the emergency room or hospitalizations due to, for example, accidents, fights, self-harm, etc.);
d. Organization of the transition process (ages of transition planning and actual passage, information sharing, family involvement, activities such as appointments and spaces to express doubts and needs);
e. In case of failed passage, the reason and the later need for care from public or private healthcare professionals;
f. Involvement of general practitioner (GP);
g. Perception of self as an adult ADHD patient, and advice that they would give to younger people about to approach transitioning age.

**Figure 1.**
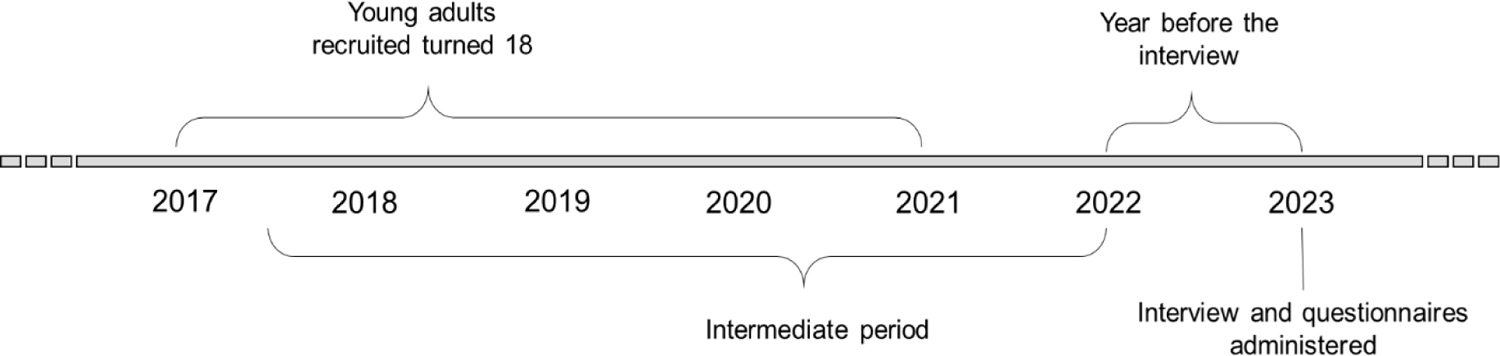
Timeline of the patients’ path. The participants of the present study turned eighteen between 2017 and 2021. They were all interviewed in 2023, and questions were related to the “last year”. Other questions referred to the time between young adults turning 18 and the year before the interview, defined as the “intermediate period.”

The questionnaires replicated questions on ADHD symptoms, their severity, and impairment areas in daily life, as well as on the organization of the transition process. Moreover, we asked whether they asked for feedback (CAMHS) or sent feedback (AMHS) on the transition’s success. The clinicians filled in the questionnaires independently, while the interviews were administered by a CAMHS worker who did not follow the patients during their care path. The questionnaires and the interview are available in the **Supplementary Material.**

Since questions included in points a, b, c, f, and g of the interviews were asked to all participants, while those in points d and e, respectively, only to referred and non-referred patients, analyses will be reported in separate blocks. The first part will describe the whole sample (e.g., clinical characteristics, occupation, daily life functioning), and the second will focus on details of non-referred and referred patients. The analyses reported are descriptive. Data are reported as the number and percentage of responders and tested using the chi-square exact test, where applicable. Median values and standard deviations summarize continuous variables.

The centers involved were ASST Santi Paolo e Carlo - San Paolo Hospital, Milan, Federico II University Hospital, Naples, and Macerata Hospital, Macerata. The ethics committee of the coordinating center, the IRCCS “Carlo Besta” Foundation Ethics Committee, approved the study (9 November 2022, protocol n. 09). All participating centers notified their ethics committees. The study followed the ethical standards of the 1964 Declaration of Helsinki and its later amendments. We followed the Strengthening the Reporting of Observational Studies in Epidemiology (STROBE) reporting guidelines.

## Results

### Descriptive characteristics of the sample

Thirty-six interviews (18 from the north, 12 from the center, and 6 from the south) were conducted, in thirteen cases with the participants’ parents (eight mothers and five fathers). The median age of the young adults at the time of the interviews was 22 years (*SD* = 1.3). According to ADHD gender prevalence (Bonati *et al*., 2021; Cortese *et al*., 2023), thirty-four males and two females participated. The flowchart (Figure 2) reports the number of patients involved in the study for the three services and the different care paths before, during, and after transition. Upon reaching adulthood, the rate of patients in care at the pediatric service differed widely between participating services (from 4 to 69%).

**Figure 2.**
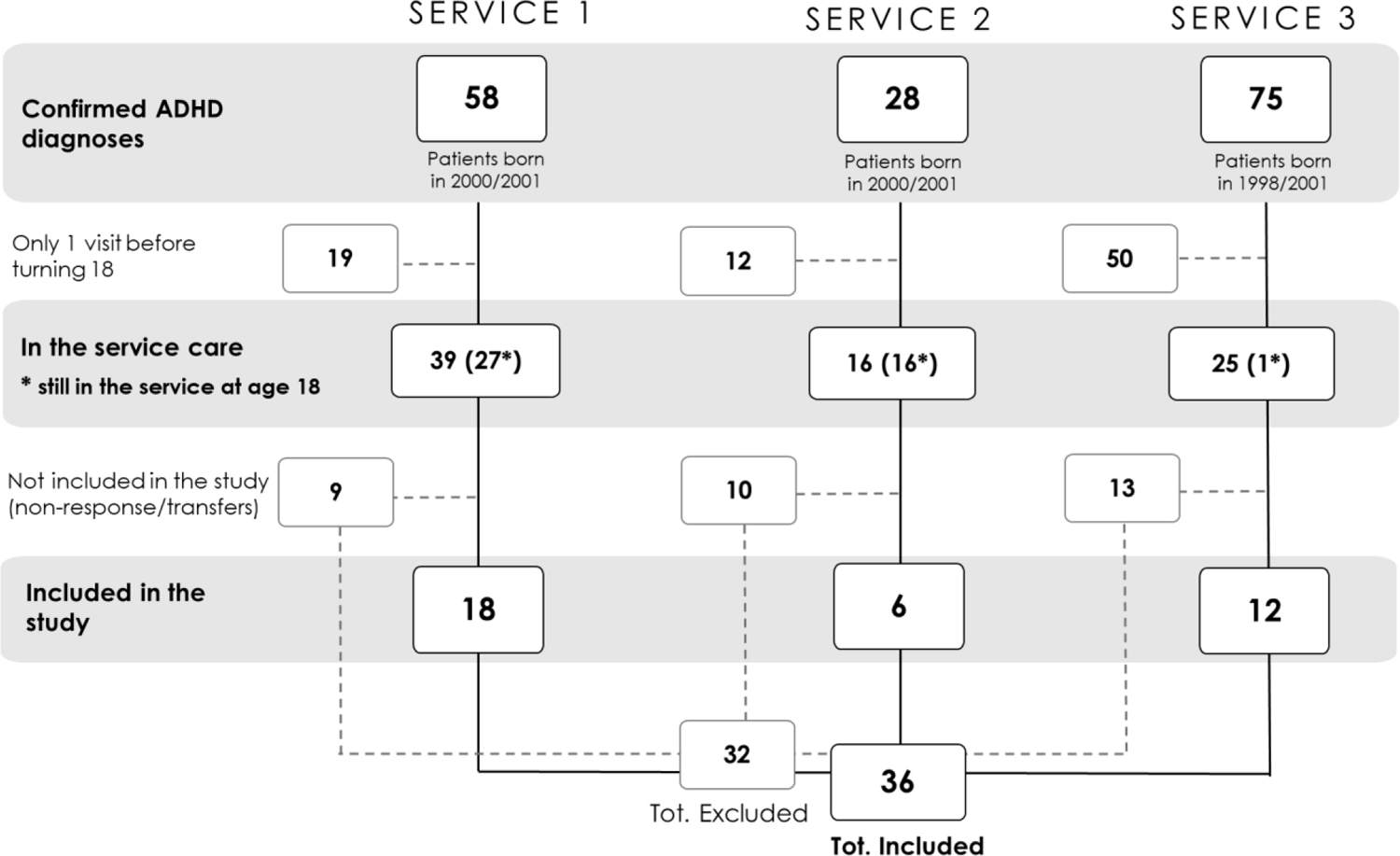
Flowchart representing the number of ADHD patients born in the target years for each service involved in the study and their continuation of care path with the service up until the study.

**At the diagnosis time**, most patients (*N*=35) had combined ADHD (i.e., with both inattentive and hyperactive-impulsive symptoms), while only one had ADHD with predominant inattention. Two-thirds of the patients received an ADHD diagnosis during primary school, while the remaining (36%) were diagnosed later. Almost all diagnoses occurred within the public healthcare system (94%), with only two cases of diagnoses in the private sector. Twenty-three patients (64%) also had one or more comorbidities: learning disorders affected 39% of patients, oppositional-provocative/behavior disorder 25%, mood disorders 8%, and autism spectrum disorder 6%. Intellectual disability, sleep disorder, and mixed specific developmental disorders were diagnosed in 3% of ADHD patients. Level of impairment **at transition** was mild in 15 cases (42%), moderate in 14 (39%), and medium/severe in 7 (19%). When turning 18, the areas of impairment most frequently reported by the CAMHS clinicians were: education/work (35 patients), emotionality (14), social/emotional relationships (13), impulsivity (11), and general quality of life (6).

Employment status **at the time of the interview** was also assessed. Seventeen young adults (47%) were working, eleven (31%) were still in high school or college, and eight were not studying and were not employed. No significant differences emerged when comparing employment between participants with mild and moderate, medium or severe impairment (Chi-square = 0.18, p > 0.99). In particular, of the fifteen with mild impairment, only three (20%) were not employed; three out of thirteen (23%) with moderate and one out of seven (14%) with medium or severe impairment were unemployed. Twenty-one participants (58%) were completely independent in daily life. The remaining 42% declared needing support with practical-organizational skills (e.g., manual dexterity, fine-motor skills, studying/ working, planning, economic and bureaucratic management, and technology use). CAMHS clinicians reported that the median duration of CAMHS stay was four years (*SD* 2.7 years; range 1-10 years), mostly (69%) since diagnosis.

Regarding **treatment**, all participants except for two received pharmacological or psychological therapy in CAMHS. Most patients (81%) received combined therapy: pharmacological and psychological (Cognitive Behavioral Therapy, Child/Parent/Teacher Training, or Psychotherapy). Some also combined different types of psychological treatment (Figure 3). Two participants underwent only psychological therapy, while three only pharmacological treatment. Methylphenidate was the medication most commonly prescribed for ADHD; thirty-one young adults (86% of the entire sample) reported taking it. Of these, twenty-four (77%) stopped it, while seven still took it when they turned 18. Atomoxetine was prescribed in the past to five patients (14% of the sample) and then stopped for all of them.

**Figure 3.**
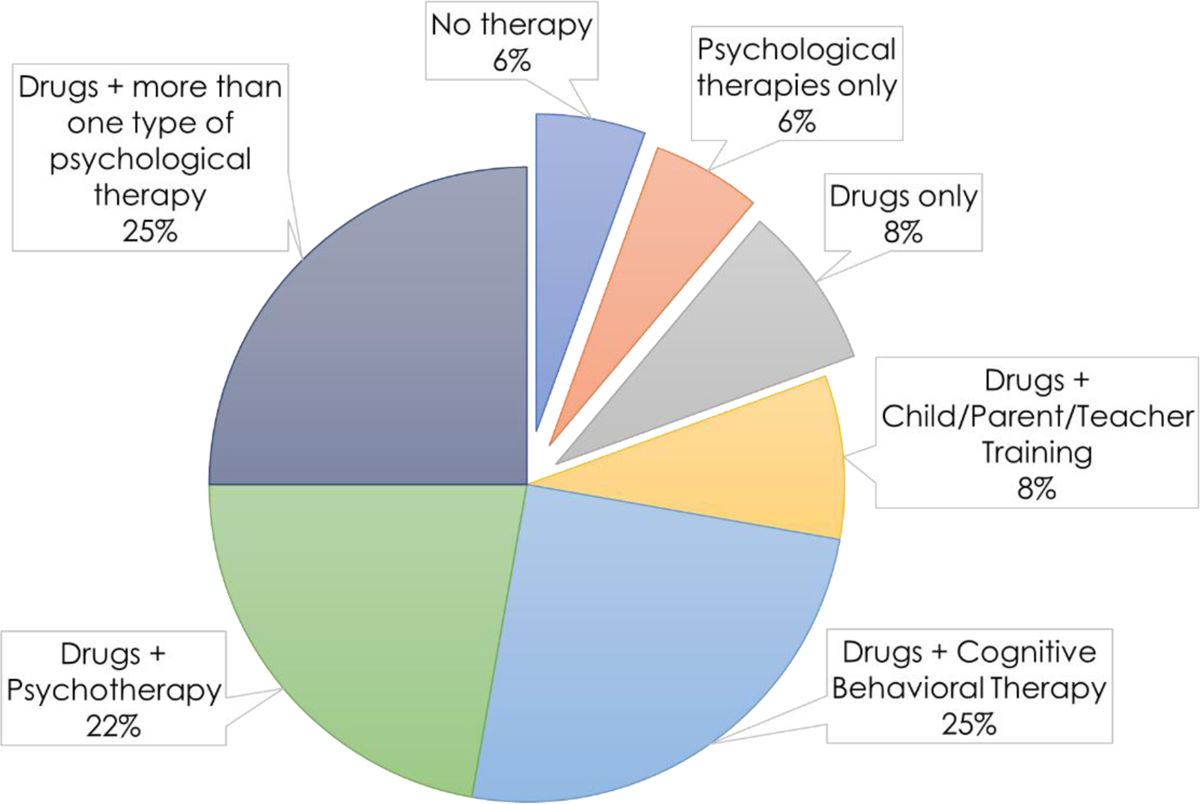
Types of treatment reported by the thirty-six young adults interviewed.

All participants were also asked about the GP’s role. Only six young adults reported that their GPs had been involved in their ADHD care path. Five said they did not need them as they preferred communicating with the specialists, but two also reported that they felt a lack of expertise for their condition. Nine did not feel the need to involve the GPs.

### Referred vs. non-referred patients

For **7 out of 36** patients, **a referral was not considered necessary** by CAMHS clinicians (one continued accessing CAMHS for medication prescriptions). Of the remaining 29 patients for whom further care was deemed necessary, a **referral to a public AMHS was not carried out for thirteen patients**. For nine of them, it was difficult to find an AMHS, and in time they were lost. Three patients dropped out of care, whereas one young adult opted for a private service. At the interview, eleven of these thirteen patients were still not in care (two attempted private care, but then stopped it), one tried to be taken into the care of an AMHS and then chose private care, and one decided autonomously to attend an AMHS due to drug addiction (and is still cared for by that service).

A **referral** to an AMHS was attempted for **sixteen** young adults. For eight of them, the referral was made **before the age of 18**, and for the other eight, when turning 18. Access to AMHS was successful in only 50% of the cases. Eight patients did not undergo a complete transition path: one was lost and then returned spontaneously to AMHS, two chose a private care path, one attempted private care but is currently not in care, one was not referred to an AMHS but transferred to a territorial CAMHS and has not changed service since, and three never attended any service again. Among the eight patients whose referral was successful, six continued their treatment at the AMHS without significant interruptions (i.e., the only six patients of the sample for which continuity of care was guaranteed), one left AMHS for an extended period and then contacted a service again, and one dropped out.

Figure 4 summarizes the paths of referred vs. non-referreds patients. A complete summary of services that cared for the patients before turning 18, in the subsequent years, and at the time of the interview, as well as some indicators of the clinical picture, is provided in the **Supplementary**

**Figure 4.**
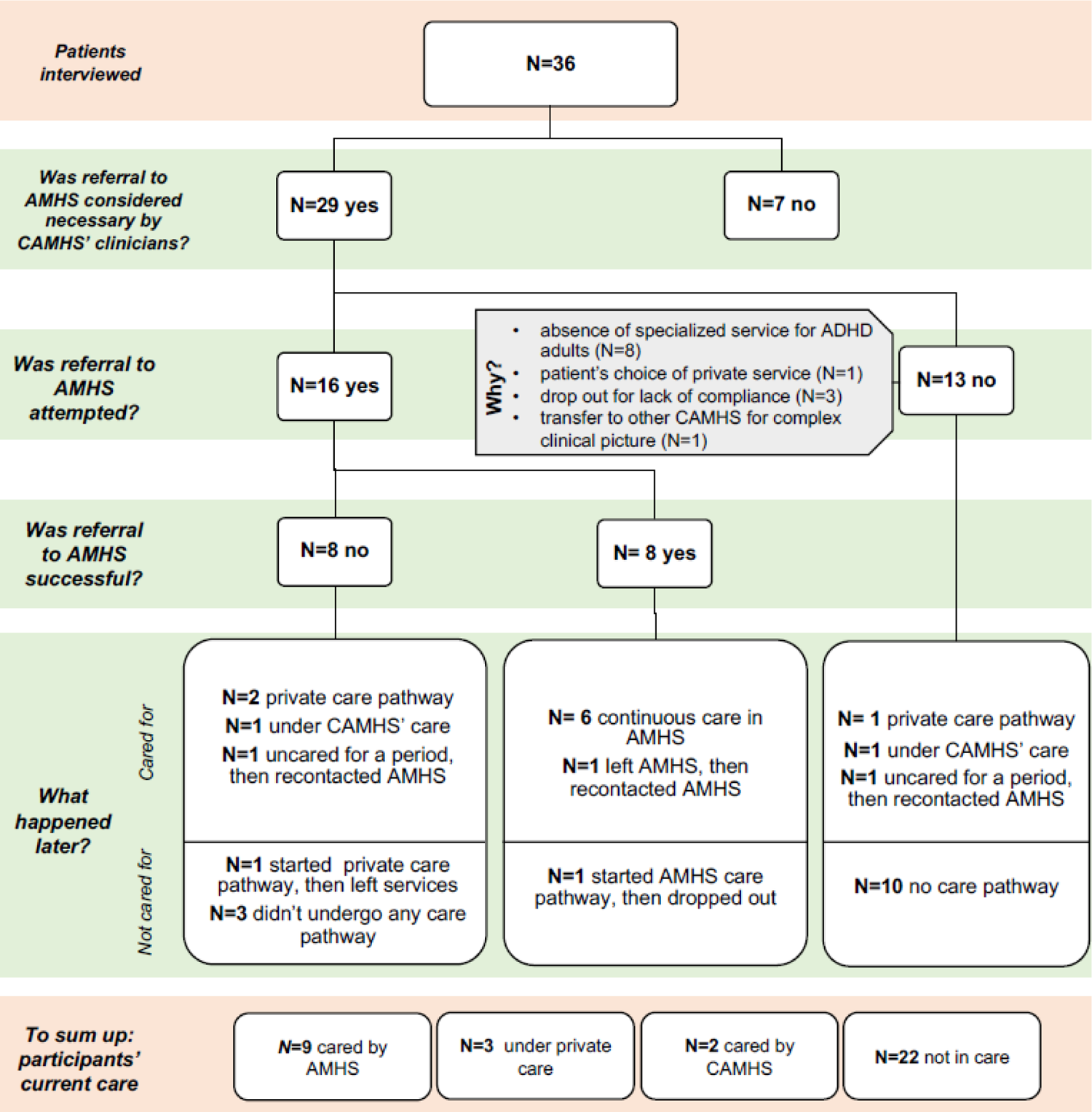
Paths of referred vs. non-referred patients.

## Material

Amongst the referred patients (*N*=16), the referral was made through informational meetings between CAMHS workers and the patient and their family in three cases. Only in one case was a case discussion between pediatric and adult services organized. For four patients, information regarding the clinical history was exchanged by a diagnostic report. The patients and their families reported significant variability between referral and assessment at the adult service: 10 months (median 10.5; SD 10.4), with a minimum of 1 month to a maximum of 36 months. Clinicians’ responses also reflected this picture, indicating a waiting time of 7 months (median 8; SD 8), with a minimum of 1 month to a maximum of 24 months. 5 CAMHS clinicians stated that more time would be needed, indicatively an additional six months.

In only one case, the CAMHS providers asked for feedback on the patient’s admission in AMHS. No follow-up meetings were organized between the services. Four of thirty-one patients continued taking methylphenidate, one was taking risperidone, and one methylphenidate and risperidone, while two were not taking any medication.

To summarize, at the interview, **twenty-seven** patients were **not cared for by a public AMHS**. They were either unsupervised (22 patients), privately cared for (3), or only afferent to CAMHS for prescription medication (2). Therefore, **nine participants** were **in AMHS care at the time of the interview** (two were also cared for by private professionals). For these nine patients, a possible referral was first addressed when they were 17 (median) and completed after they turned 18 (median; SD 0.93). Five of the **nine participants in AMHS care at the time of the interview** had moderate, two medium/severe, and two mild levels of impairment. Only one reported sentinel events in the year preceding the interview due to substance abuse.

### Well-being indicators

At the interview, two out of seven adolescents with a medium/severe **impairment** were cared for by an AMHS, while three were cared for by a private mental health professional, and two dropped out. Eight out of fourteen adolescents with a moderate impairment were not cared for by a healthcare service (four of whom required support for daily life activities and/or presented sentinel events), five attended an AMHS, and one remained in a CAMHS.

Eight patients presented at least one **sentinel event**, and six were never referred (even if their clinicians evaluated half of them as needing further care). Of the twenty-seven young adults not currently cared for by an AMHS (seventeen with good general well-being, ten with marked problems in various areas of life), in the year preceding the interview, one case of emergency room admission following fights, one follow-up examination after a drunk driving episode, one substance abuse, and four episodes of car accidents were reported.

Nineteen patients reported a need for support in daily life activities and/or a sentinel event in the year before the interview: 8 sought a mental health professional (6 AMHS, 2 private), while eleven were unsupervised. **Eleven individuals with a high need for care** could be identified based on the degree of impairment (moderate or medium/severe) associated with support needs or sentinel events. Five of these were not followed by a mental health professional at the time of the interview. Moreover, three were among those patients for whom a referral was deemed unnecessary.

When comparing the above indicators, it can be observed that the level of impairment and the proportion of young adults needing support for daily life activities is higher in patients attending an AMHS and in those with an unsuccessful referral. On the contrary, the rate of sentinel events is greater in adults not referred to an AMHS (Table 1). The proportions of the indicators is similar in those with a successful versus unsuccessful referral.

**Table 1.** Number of patients presenting level of impairment moderate or medium/severe, need for support, presence of sentinel events and unemployment. These patients are grouped into those for which a referral was successful, unsuccessful, or not carried out (respectively, 9, 7, and 20 in the full sample).

|  | AMHS (N=9) | Unsuccessful (N=7) | Not referred (N=20) |
| --- | --- | --- | --- |
| moderate or medium/severe impairment | 7 | 5 | 9 |
| need for support (yes) | 5 | 3 | 7 |
| sentinel events (yes) | 1 | 1 | 6 |
| unemployment | 5 | 1 | 2 |

### Acquired skills, specific experiences

All thirty-six young adults were asked questions to explore their perception of skills acquisition during the transition. Almost all (eight out of nine) of the patients currently in AMHS care reported not having acquired any particular skills. In contrast, a similar perception for those not in care was present for only 37% (10 participants). As for the description of their transition experience, unassisted young adults and those under AMHS care reported very similar observations. For instance, they found it difficult to handle everyday life, especially in organizational aspects and social exchanges, with difficulty managing anger and impulsiveness. They agreed on the importance of getting help from close people and trusting doctors. While they also agreed that in time, it became easier to learn how to manage symptoms, only young adults still in AMHS care stressed the importance of taking medications and preparing for the passage to adult services. On the other hand, the unassisted young adults reported greater difficulties in the school/work environment and marked as desirable more practical strategies and support from teachers.

## Discussion

Clinics dedicated to transition were developed in France (Le Roux *et al*., 2023), whereas elsewhere, programs such as those of the Got Transition Six Core Elements in the United States, the NICE Transition Guidelines, and Ready Steady Go in the United Kingdom (van Staa, Peeters and Sattoe, 2020) were set up. However, even if research and innovative practices in transitional care for young people with chronic conditions have improved in the last few years, barriers remain. Many people experience discontinuity of care and drop out during this critical period (Reneses *et al*., 2023). Inadequate transition processes not only pose risks to mental health, but can also exacerbate the likelihood of engaging in at-risk behaviors. They may even contribute to inequalities in care, which every individual should be guaranteed as a fundamental human right (Munyikwa *et al*., 2023). Supporting patients with ADHD during transitions is crucial, and clinicians must be empowered to facilitate an appropriate transition process (Scarpellini and Bonati, 2022).

The results here reported indicate the urgency of timely interventions to facilitate the transition of ADHD patients from pediatric to adult services. This successful transition rate is similar to that described in other countries (Eke *et al*., 2020; Maurice *et al*., 2022), but still inadequate. Low referral acceptance and lack of communication between CAHMS and AMHS contribute to the loss of care for approximately half of patients. Such mechanisms result from scarce services and resources, as previously highlighted elsewhere (Eke *et al*., 2020; Appleton *et al*., 2023; Roberti *et al*., 2023b).

The first result of the present study is that a referral to an AMHS was carried out in nearly half of the cases. Often, patients were not referred since the CAMHS clinicians evaluated the level of impairment as mild. Nonetheless, for a non-negligible number of cases, it was challenging to find a service that could care for adult patients. The situation is even worse when considering that only one out of four young patients deemed in need of continued treatment by psychiatrists were successfully transferred to AMHS. Moreover, it is worrying that only one out of three patients with medium/severe impairment and nearly half of the subjects with a potentially high level of care needs attended AMHS at the time of the interview.

The second main finding is that transition paths are related to the type of child psychiatric service (Figure 2). While the most common scenario in a university hospital (service 2) is an early referral of patients, most patients are referred at 18 years of age in a territorial structure associated with a university (service 1). On the other hand, in service 3, continuity of care appears to be difficult as patients are lost even before turning 18. Nonetheless, 33% of the patients were described as still having a medium impairment level, and for 25% of them, a referral to adult services would have been necessary. This difference between services may be due to each hospital’s different networks and resources. In the future, care policies should address these differences.

When viewing the transition pathways, a few patients had already been transferred to AMHS when they turned 18, while more were transferred in the following years. Notably, only nine were still in care at the interview. The proportion of referred patients and successful referrals is the same when restricting the analysis to the homogeneous sample of patients who were still in the CAMHS at 18 years of age, suggesting that our findings are scantly influenced by the dropout of the adolescents from CAMHS before 18 years of age.

The dropout from the AMHS is an inconvenience of the treatment path that must be carefully evaluated considering the possible outcomes. In fact, of the nine patients cared for by an AMHS, six had a continuous care path, while three were lost in the intermediate period and then returned following a worsening of symptoms. Two of them reported having developed drug addiction in the meantime. They described their experience as follows: “…We ADHD kids are at risk with drugs because we physiologically need to feel strong emotions. If we’re not well aware and prepared, it’s a snap to ‘get into it’, try it even once, and it’s too late. I realized this to my own expense, unfortunately”, and “It is important not to disengage from services after age 18; I would advise against making the same mistake I made. As adults, we feel like ‘heroes’ who can do and decide everything on our own, but it is very easy to end up in bad situations”.

Other sentinel events were highlighted in the study, i.e., critical episodes that lead to emergency room access in the healthcare system, recurrent relational difficulties, reactivity and impulsivity, attentional difficulties, anxious symptomatology, as well as being unemployed. Unemployment also seemed common among those who were not referred to an AMHS, but would have needed it. As already described, the risk of unemployment in ADHD young adults is high, even up to 70% (Helgesson *et al*., 2023). Episodes of fights, drunk driving episodes, and car accidents must not go unnoticed. In our sample, these episodes were reported in 88% of the cases by these patients who were not in AMHS care. It follows that it is essential that warning signs are promptly ascertained and reported whenever they appear. Therefore, GPs (who seem not currently involved in ADHD management) should be encouraged to advise their patients to resume a specific care path should they detect these signs. The GP’s role can be particularly relevant when considering that half of the patients with a high need for care were not cared for by a mental health professional.

Lastly, considering that universal health care is guaranteed in Italy, it is striking that one in five patients felt the need to contact a private mental health professional at least once during the care pathway, particularly after leaving the CAMHS.

It should be kept in mind that the description provided by the present study is limited to a few Italian centers and that the sample was recruited voluntarily and could be biased. We, however, managed to represent a variety of situations from independence in daily life (present for 58% of young adults) to drugs and therapies administered, and several types of paths and clinical case complexities, i.e., the real world. The emerging figure is worrying, although not exclusive to Italy, and should encourage us to promptly undertake improvement initiatives (Swift *et al*., 2013; Leavey *et al*., 2019). It is crucial to further explore the factors that lead to unsuccessful referrals in the future. Other than the lack of resources, dedicated specialized public services, and follow-up, the transition period per se should also be analyzed carefully. The TransiDEA project’s third phase will aim to design a process that begins earlier and is participatory among the different providers involved, the young adults, and their families. This will allow us to jointly define guidelines for a more appropriate transition pathway by involving clinicians, young adults, scientific societies, and patient associations.

While the defined consensus document will be drafted based on the experiences of young adults and services in Italy, we hope that it will encourage other nations to replicate a similar model.

## List of Abbreviations

ADHD: Attention Deficit/Hyperactivity Disorder

CAMHS: Child and Adolescent Mental Health Services

AMHS: Adult Mental Health Services

GP: general practitioner

NICE: National Institute for Clinical Health and Excellence

TransiDEA: Transition in Diabetes, Epilepsy, and ADHD patients.

## Declarations

### Ethical standards

The authors assert that all procedures contributing to this work comply with the ethical standards of the relevant national and institutional committees on human experimentation and with the Helsinki Declaration of 1975, as revised in 2000. The study is part of a wider project (”*Transition care between adolescent and adult services for young people with chronic health needs in Italy”, RF-2019-12371228*) that was approved by the IRCCS “Carlo Besta” Ethics Committee (ethics committee of reference for the Mario Negri IRCCS Institute) (8 September 2021, protocol n. 87). The present study as part 2 of the project was approved by the IRCCS “Carlo Besta” Ethics Committee (9 November 2022, protocol n. 09) and notified to all the involved centers’ Ethics Committees. All participants signed an informed consent prior to their participation.

### Availability of data and materials

The data presented in this study are available on the Zenodo platform (https://zenodo.org/doi/10.5281/zenodo.10843581). The materials used (structured interview and questionnaires) are available online as supplementary material.

## Acknowledgments

We would like to thank the TransiDEA Group members: ***Coordinating and Managing Group***: Istituto di Ricerche Farmacologiche Mario Negri IRCCS, Milan, Italy: Maurizio Bonati, Antonio Clavenna, Francesca Scarpellini, Elisa Roberti, Rita Campi, Massimo Cartabia, Michele Giardino, Michele Zanetti, Maria Grazia Calati; ***Diabetes (D):*** AUSL della Romagna, Ravenna, Italy: Vanna Graziani, Federico Marchetti, Tosca Suprani; Santa Maria delle Croci Hospital, Ravenna, Italy: Paolo Di Bartolo; ***Epilepsy (E):*** ASST Santi Paolo e Carlo – Ospedale San Paolo, Milano: Maria Paola Canevini, Ilaria Viganò; ***ADHD (A):*** ASST Santi Paolo e Carlo – Ospedale San Paolo, Milano: Ilaria Costantino, Valeria Tessarollo; University of Milan, Milan, Italy: Eleonora Basso. The authors would also like to acknowledge Chiara Pandolfini for language editing.

## Author Contributions

MB conceptualized the study with the help of the TransiDEA Group; MB, AC and FS curated methodology; RC, MG, and MZ curated resources; CB, MPR, EB, IC, VT, MP, MD, and CG contributed to the collection of information through interviews; EB and ER carried out a primary data analysis; ER and MB wrote the original manuscript draft; ER, MB, FS and AC reviewed and edited the manuscript. MB supervised the project. All authors have read and agreed to publish the current version of the manuscript.

