## Supplementary Material for "The Transition from adolescence to adulthood mental health services for young people with ADHD in Italy"

### Appendix

#### 1. Transition process assessment - questionnaire for ADHD patients

*Structured interview for patients* *<u>who turned 18 years of age</u>* *between 2018 and 2019.*

*The person administering the interview must not be the practitioner who directly followed the boy/girl during the transition process. The interview can be administered to the patient together with the parent/caregiver. If the respondent is not the patient but a family member/caregiver, please ask them to answer the questions with the patient's care path in mind.*

*Concerning the questions where "day, month, year" is asked to be entered, it is also okay to complete only the month and year information if the respondent does not remember the exact day.*

##### 1.1 - Version for patients sent to an AMHS

Answers are provided by:
❑ Patient
❑ Family member, please specify: ______________________________

Patient's date of birth: day |__|__|, month |__|__|, year |__|__|__|__|

Gender of the patient: M❑ F ❑

Date you were notified of the diagnosis: day |__|__|, month |__|__|, year |__|__|__|__|
At:
❑ Hospital or CAMHS - name: _________________________
❑ Private service - specify discipline (e.g., psychologist): ________________________________

Have you received sufficient information about the diagnosis? ❑ YES ❑ NO

Have you received any other diagnosis besides ADHD?

___________________________ ___________________________

**1**. What ADHD treatment medications have you taken to date?

❑ Methylphenidate Start date: day |__|__|, month |__|__|, year |__|__|__|__|

Ongoing treatment? ❑ YES ❑ NO

If NO, date of discontinuation: day |__|__|, month |__|__|, year |__|__|__|__|

❑ Atomoxetine Start date: day |__|__|, month |__|__|, year |__|__|__|__|

Ongoing treatment? ❑ YES ❑ NO

If NO, date of discontinuation: day |__|__|, month |__|__|, year |__|__|__|__|

❑ Other – specify which ones and why: ____________________________________________

Start date: day |__|__|, month |__|__|, year |__|__|__|__|

Ongoing treatment? ❑ YES ❑ NO

If NO, date of discontinuation: day |__|__|, month |__|__|, year |__|__|__|__|

Have you experienced any side effects after taking any of these drugs?

_____________________________________________________________________________________

**2**. What psychological therapy for ADHD have you received from the time of diagnosis to the present?

❑ Cognitive-behavioral therapy (includes psychoeducation, coaching, and mindfulness)

Start date: day |__|__|, month |__|__|, year |__|__|__|__|

Ongoing treatment? ❑ YES ❑ NO

If NO, date of discontinuation: day |__|__|, month |__|__|, year |__|__|__|__|

❑ Child training, or Parent training (aimed at parents) or Teacher training (aimed at teachers)

Specify: ______________________________________

Start date: day |__|__|, month |__|__|, year |__|__|__|__|

Ongoing treatment? ❑ YES ❑ NO

If NO, date of discontinuation: day |__|__|, month |__|__|, year |__|__|__|__|

❑ Psychotherapy - specify type (Examples: Schema Therapy, Systemic-Relational Therapy, Psychodynamic Therapy): ______________________________________________

Start date: day |__|__|, month |__|__|, year |__|__|__|__|

Ongoing treatment? ❑ YES ❑ NO

If NO, date of discontinuation: day |__|__|, month |__|__|, year |__|__|__|__|

❑ Other - specify: ______________________________________________

Start date: day |__|__|, month |__|__|, year |__|__|__|__|

Ongoing treatment? ❑ YES ❑ NO

If NO, date of discontinuation: day |__|__|, month |__|__|, year |__|__|__|__|

**3.** Did you perceive the treatment as appropriate for your situation? Would you have liked to have received another type of treatment? If yes, which one?

______________________________________________________________________________

**4**. Before turning 18, were you followed by other services besides CAMHS for the treatment of ADHD?

❑ YES - please indicate which: __________________________________________

❑ NO

**5**. When you turned 18, by which service(s) or specialist(s) were you followed? (more than one option can be selected)

❑ CAMHS

❑ AMHS

❑ General practitioner

❑ Private psychiatrist/psychologist/psychotherapist/private center

❑ Other - specify: ________________________________________________________________

**6**. Since you turned 18 to the present, have you been followed by other practitioners/services?

❑ YES

❑ NO

If YES, which one(s) (more than one option can be selected)?

❑ CAMHS

❑ AMHS

❑ General practitioner

❑ Private psychiatrist/psychologist/psychotherapist

❑ Other - specify: ________________________________________________________________

**7.** Currently:

- ❑ I am completely independent in all daily activities
- ❑ I need help with some daily activities - specify: ___________________________
- ❑ I need help with most daily activities

**8.** School and work:

Are you currently attending any school or college?

- ❑ YES: school/professional courses/university - specify: _______________________________
- ❑ NO: I quit at the age of: ________________________________________________________
  reason: _________________________________________________________________

Highest educational title: _____________________________________________________________

Are you currently working?

- ❑ YES - specify in what area: _________________________________
- ❑ NO

**9**. Who do you currently live with?

- ❑ Alone
- ❑ Partner
- ❑ Parent(s)/caregiver(s)
- ❑ Roommate(s)

**10. What service(s) or provider(s) are you currently followed by?** (more than one option can be selected)

- ❑ CAMHS
- ❑ General practitioner
- ❑ Private psychiatrist/psychologist/psychotherapist/private center
- ❑ AMHS
  - ➔ *Fill out SECTION A - Patients followed by adult services*
- ❑ None
  - ➔ *Fill out SECTION B -Patients not followed by any service*
- ❑ Other - specify: _______________________________________________________________

**<u>SECTION A - Patients followed by adult services.</u>**

**1a.** Currently, which adult mental health service is following you?

____________________________________________________________

**2a.** During the past year, have you made any outpatient visits (for any needs, excluding ADHD)?

- ❑ YES - specify for what: __________________________________________________________
- ❑ NO

**3a**. In the past year, have you made any day-hospital visits (for any needs, excluding ADHD)?

- ❑ YES - specify for what: __________________________________________________________
- ❑ NO

**4a**. In the past year, have you had any hospitalizations (for any need, excluding ADHD)?

- ❑ YES - specify for what: __________________________________________________________
- ❑ NO

***Transition preparation phase:***

**5a.** When was the subject of your transfer to the psychiatric service first broached? How old were you?

__________________________________________________________________________________

**6a.** Has CAMHS actively participated in the referral to Mental Health services for adults in transition?

- ❑ YES
- ❑ NO

**7a.** Was there a preparation phase Before sending you to the current service?

- ❑ YES
- ❑ NO

If YES, who planned the transition process?

- ❑ CAMHS
- ❑ Other figure - specify: ________________________________

If YES, how old were you? ______

If YES, did you acquire new skills at this stage?

- ❑ YES
- ❑ NO

If YES, which ones (more than one option can be selected)?

- ❑ Knowledge of the disorder (onset, symptoms, persistence)

❑ Knowledge of comorbidities (possible other conditions concomitant to ADHD)

❑ Knowledge of primary treatments (psychological, educational, pharmacological)

❑ Knowledge of drugs (side effects, dosage)

❑ How to book appointments with the service (who to contact, what materials to bring)

**8a.** Which figures were involved in the preparation phase? (more than one option can be selected)

❑ Physician

❑ Psychologist

❑ Nurse

❑ Pediatrician/general practitioner

❑ Educator

❑ Other (specificare) ________________________________________________

**9a.** did you have a chance to express your doubts and concerns during transition?

❑ YES - which ones?

________________________________________________________________________________

❑ NO - why?

________________________________________________________________________________

**10a.** Have your concerns been addressed**?**

❑ YES

❑ NO

**11a**. Have you been informed about the modalities of the transition process?

❑ YES

❑ NO

**12a**. Have you been informed about the timing of the transition process?

❑ YES

❑ NO

**13a**. Have you been informed about the reasons for the transition process?

❑ YES

❑ NO

**14a**. Did the CAMHS involve you and your family in preparing for the transfer of care?

❑ YES - as: __________________________________________________________________

❑ NO

**15a. Are there** questions you were addressed but were not? Or are there questions that were not answered?

______________________________________________________________________________

***Transfer phase***

**16a**. How old were you at the time of transfer? _____

**17a.** How did the transfer contact take place?

❑ The AMHS contacted you

❑ You received a letter

❑ Your parents received a letter

❑ Your referring clinician at the CAMHS has contacted the AMHS

❑ You actively contacted the AMHS

❑ Other (specify) ___________________________________________________________

**18a.** Do you think there have been difficulties in finding an appropriate adult service for you?

❑ YES (specify)_____________________________________________________________

❑ NO

**19a.** Activities carried out during transfer:

- Interviews inherent to the path to be taken:

  ❑ YES ❑ NO
- Discussions with your parents inherent to the path to be taken:

  ❑ YES ❑ NO
- Meetings between the two Services (CAMHS and adult service):

  ❑ YES ❑ NO
- Interview exclusively with the adult service psychiatrist:

  ❑ YES ❑ NO
- Involvement of others (general practitioner, psychologist, nurse, etc.):
- ❑ YES - specify: ____________________ ❑ NO
- Involvement of other professionals (educators, curricular or support teachers, social services, educators, etc.):

  ❑ YES - specify: ____________________❑ NO

**20a.** Was there a time when you were followed by the two services (CAMHS and AMHS) simultaneously?

❑ NO

❑ YES, duration: |___|___| (months); number of joint appointments: |___|___|

**21a.** How many meetings were conducted during the dispatch phase? _______

How often (how many times a week)? _______

**22a.** In which mode?

❑ Presence

❑ Online

❑ Mixed form

**23a.** How much time passed between referral and assessment at the adult service? __________(months)

**24a**. While waiting:

❑ You were aware of what was going to happen

❑ It was clear to whom you could turn for help or support

❑ Did you have a phone number / could you still contact the CAMHS

❑ Did you have a phone number / could you contact the AMHS

❑ You contacted/attempted to contact someone

**25a**. In the meantime, has your drug treatment continued smoothly?

❑ YES

❑ NO - Why? _______________________________________________________

❑ At that time, I was not taking medication

**26a**. Has your drug treatment been changed since then?

❑ YES - How? _______________________________________________________

❑ NO

**27a.** At the first appointment with the psychiatric service**:**

❑ You knew who he would meet and where to go

❑ You did not know who he would meet and where to go

**28a.** How do you think the two services worked together?

bad sufficiently well very well

In your opinion, what is the level of quality of care received?

low sufficient good high

In your opinion, what is the quality level of the transition process carried out?

low sufficient good high

In your opinion, how useful were the activities carried out during the transition process?

not useful sufficiently useful useful very useful

***Role of the general practitioner***

Has your primary care physician been involved in ADHD treatment so far?

- ❑ YES
- ❑ NO

If you needed to, did you contact him? For what reasons? If you did not contact him, why?

___

Do you think the general practitioner can help you in any way?

___

**Taking care of oneself**

Now that you have moved to adult services, do you think you have become more autonomous with respect to managing ADHD? For example, making appointments, ordering medication, and going to appointments. (If they mention the mother/father, ask "if your mom wasn't doing all these things, would you know how to do them yourself?")

___

Is there anything specific that you find particularly difficult to manage as an adult with ADHD?

___

***Final questions.***

If you were to talk to a younger person with ADHD still being treated at the CAMHS, what would you tell him to expect once he becomes an adult?

___

What advice would you give him about the type of treatment, symptoms, medications?

___

What advice would you give him about transferring to adult services?

___

In your opinion, what was missed or needed to be done differently during your journey?

___

**SECTION B - Patients not followed by any service.**

**1b.** Why is it not currently followed by any service?

- ❑ I don't feel I need it
- ❑ I could not find a service that met my needs, so I gave up
- ❑ I could not find a service that met my needs but I am still looking for
- ❑ I am waiting to be contacted

**2b.** In the past years (from when you turned 18 to the present), have you been followed by one or more services?

- ❑ YES
- ❑ NO

If YES, by whom?

- ❑ CAMHS
- ❑ AMHS
- ❑ General practitioner
- ❑ Private psychiatrist/psychologist/psychotherapist

If YES, why did you stop using this service(s)?

___

___

**3b.** During the past year (as of today), have you made any outpatient visits (for any needs, excluding ADHD)?

- ❑ YES - specify for what: ___
- ❑ NO

**4b**. In the past year, have you made any day-hospital visits (for any needs, excluding ADHD)?

- ❑ YES - specify for what: ___

- ❑ NO

**5b.** In the past year, have you had any hospitalizations (for any need, excluding ADHD)?

- ❑ YES - specify for what: ____________________________________________
- ❑ NO

**6b**. Do you feel you have acquired skills during the years since you turned 18?

- ❑ YES
- ❑ NO

If YES, which ones:

- ❑ Knowledge of the disorder (onset, symptoms, persistence)
- ❑ Knowledge of comorbidities (possible other conditions concomitant to ADHD)
- ❑ Knowledge of primary treatments (psychological, educational, pharmacological)
- ❑ Knowledge of drugs (side effects, dosage)
- ❑ How to book appointments with the service (who to contact, what materials to bring)

***Role of the general practitioner***

Has your primary care physician been involved in ADHD treatment so far?

- ❑ YES
- ❑ NO

If you needed, did you contact him? For what reasons? If you did not contact him, why?

______________________________________________________________________

Do you think the general practitioner can help you in any way?

______________________________________________________________________

**Taking care of oneself**

Over the years, because of the support you have received, do you think you have become more independent concerning managing ADHD? For example, making appointments, ordering medication, and going to appointments. (If they mention the mother/father, ask "If your mom didn't do all these things, would you know how to do them yourself?")

______________________________________________________________________

Is there anything specific that you find particularly difficult to manage as an adult with ADHD?

______________________________________________________________________

***Final questions.***

If you were to talk to a younger person with ADHD still being treated at the CAMHS, what would you tell him to expect once he becomes an adult?

---

What advice would you give him about the type of treatment, symptoms, medications?

---

In your opinion, what was missed or needed to be done differently during your journey?

---

#### 1.2 - Version for patients who remained in CAMHS

Answers are provided by:

❑ Patient

❑ Family member, please specify: ______________________________

Patient's date of birth: day |__|__|, month |__|__|, year |__|__|__|__|

Gender of the patient: M❑ F ❑

Date you were notified of the diagnosis: day |__|__|, month |__|__|, year |__|__|__|__|

At:

❑ Hospital or CAMHS - name: _________________________

❑ Private service - specify discipline (e.g., psychologist): ________________________________

Have you received sufficient information about the diagnosis? ❑ YES ❑ NO

Have you received any other diagnosis besides ADHD?

___________________________ ___________________________

**1**. What ADHD treatment medications have you taken to date?

❑ Methylphenidate Start date: day |__|__|, month |__|__|, year |__|__|__|__|

Ongoing treatment? ❑ YES ❑ NO

If NO, date of discontinuation: day |__|__|, month |__|__|, year |__|__|__|__|

❑ Atomoxetine Start date: day |__|__|, month |__|__|, year |__|__|__|__|

Ongoing treatment? ❑ YES ❑ NO

If NO, date of discontinuation: day |__|__|, month |__|__|, year |__|__|__|__|

❑ Other – specify reason: ___________________________________________

Start date: day |__|__|, month |__|__|, year |__|__|__|__|

Ongoing treatment? ❑ YES ❑ NO

If NO, date of discontinuation: day |__|__|, month |__|__|, year |__|__|__|__|

Have you experienced any side effects after taking any of these drugs?

________________________________________________________________________

**2**. What psychological therapy for ADHD have you received from the time of diagnosis to the present?

❑ Cognitive-behavioral therapy (includes psychoeducation, coaching, and mindfulness)

Start date: day |__|__|, month |__|__|, year |__|__|__|__|

Ongoing treatment? ❑ YES ❑ NO

If NO, date of discontinuation: day |__|__|, month |__|__|, year |__|__|__|__|

❑ Child training, or Parent training (aimed at parents) or Teacher training (aimed at teachers)

Specify: ______________________________

Start date: day |__|__|, month |__|__|, year |__|__|__|__|

Ongoing treatment? ❑ YES ❑ NO

If NO, date of discontinuation: day |__|__|, month |__|__|, year |__|__|__|__|

❑ Psychotherapy - specify type (Examples: Schema Therapy, Systemic-Relational Therapy, Psychodynamic Therapy): ____________________________________

Start date: day |__|__|, month |__|__|, year |__|__|__|__|

Ongoing treatment? ❑ YES ❑ NO

If NO, date of discontinuation: day |__|__|, month |__|__|, year |__|__|__|__|

❑ Other - specify: ____________________________________

Start date: day |__|__|, month |__|__|, year |__|__|__|__|

Ongoing treatment? ❑ YES ❑ NO

If NO, date of discontinuation: day |__|__|, month |__|__|, year |__|__|__|__|

**3.** Did you perceive the treatment as appropriate for your situation? Would you have liked to have received another type of treatment? If yes, which one?

________________________________________________________________________

**4**. Before turning 18, were you followed by other services besides the CAMHS for ADHD treatment?

❑ YES - please indicate which ones: ________________________________

❑ NO

**5**. At the time you turned 18, by which service(s) or specialist(s) were you being followed? (more than one option can be selected)

❑ CAMHS

❑ AMHS

❑ General practitioner

❑ Private psychiatrist/psychologist/psychotherapist/private center

❑ Other - specify: ____________________________________________________

**6**. Since you turned 18 to the present, have you been followed by other practitioners/services?

❑ YES

❑ NO

If YES, which one(s) (more than one option can be selected)?

❑ CAMHS

❑ AMHS

❑ General practitioner

❑ Private psychiatrist/psychologist/psychotherapist

❑ Other - specify: ____________________________________________________

**7.** What service(s) or provider(s) are you currently followed by? (more than one option can be selected)

❑ CAMHS

❑ AMHS

❑ General practitioner

❑ Private psychiatrist/psychologist/psychotherapist/private center

❑ None

❑ Other - specify: ____________________________________________________

**8.** Currently:

❑ I am completely independent in all daily activities

❑ I need help with some daily activities - specify: ___________________________

❑ I need help with most daily activities

**9.** School and work:

Are you currently attending any school or college?

❑ YES: school/professional courses/university - specify: ______________________________

❑ NO: I quit at the age of: ____________________________________________________

why: ____________________________________________________

Highest educational title: ____________________________________________________

Are you currently working?

❑ YES - specify in what area: _________________________________

❑ NO

**10**. Who do you currently live with?

- ❑ Alone
- ❑ Partner
- ❑ Parent(s)/caregiver(s)
- ❑ Roommate(s)

**Questions about current care**

**1a.** When was your last appointment at CAMHS? day |__|__|, month |__|__|, year |__|__|__|__|

**2a.** During the past year, have you made any outpatient visits (for any needs, excluding ADHD)?

- ❑ YES - specify for what: ________________________________________________
- ❑ NO

**3a**. In the past year, have you made any day-hospital visits (for any needs, excluding ADHD)?

- ❑ YES - specify for what: ________________________________________________
- ❑ NO

**4a**. In the past year, have you had any hospitalizations (for any need, excluding ADHD)?

- ❑ YES - specify for what: ________________________________________________
- ❑ NO

**5a**. Since reaching adulthood, have you perceived any differences concerning the frequency of appointments at CAMHS and/or the type of activities and treatments offered by the service (refer to the ADHD pathway)

- ❑ YES - such as: ____________________________________________________________
- ❑ NO

**6a.** In your opinion, why are you still in the CAMHS care after turning 18?

________________________________________________________________________________

#### A. ROLE OF THE GENERAL PRACTITIONER

Has your primary care physician been involved in ADHD treatment so far?

- ❑ YES
- ❑ NO

If you needed, did you contact him? For what reasons? If you did not contact him, why?

---

Do you think the general practitioner can help you in any way?

---

#### B. TAKE CARE OF ONESELF

Now that you are older, do you think you have become more independent concerning managing ADHD? For example, making appointments, ordering medication, and going to appointments. (If they mention his mother/father, ask, "If his mom wasn't doing all these things, would he know how to do them himself?")

---

Is there anything specific that you find particularly difficult to manage as an adult with ADHD?

---

#### C. FINAL QUESTIONS.

If you were to talk to a younger person, what would you tell him to expect once he becomes an adult?

---

What advice would you give him about the type of treatment, symptoms, medications?

---

In your opinion, what was missed or needed to be done differently during your journey?

---

#### 2. Transition process assessment - questionnaire for ADHD patients' clinicians

##### 2.1 - Version for CAMHS clinicians

*We kindly ask you to fill out this questionnaire for each of the ADHD patients you have personally followed when they turned 18 and when they may have been transferred (transitioned) from a Child Neuropsychiatry Service (CAMHS) to an adult service (AMHS). These patients should have turned 18 between 2018 and 2019 and should currently be between 20 and 24 years old. The person filling out the questionnaire must be the practitioner who directly followed the patient.*

The questionnaire is completed by (professional figure): ________________________________

Compiler's email: _________________________________

Patient's date of birth: day |__|__| month |__|__| year |__|__|__|__|

Gender of the patient: M❑ F ❑

Patient diagnosis:

❑ ADHD - Predominant Inattention

❑ ADHD - Predominant hyperactivity/impulsivity

❑ ADHD - Combined

Indicate in which areas was present an impairment at the time of transition:

❑ Education/work

❑ Social/emotional relationships

❑ Impulsivity (e.g., money management, gambling, substance use, accidents/risk behaviors, sexuality)

❑ Emotionality (depression, anxiety, obsessions, fears/worries)

❑ Quality of life (sleep, nutrition, physical activity)

Indicate the patient's level of impairment at the time of transition:

❑ Mild

❑ Moderate

❑ Medium-severe

❑ Severe

Drug therapy when turning 18:

❑ Yes - which one: ________________________________________________________

❑ NO

How long has the ADHD patient been in the center's care?

❑ Since the diagnosis

❑ Subsequent care (specify for how many years the patient was followed: ____________)

Has the patient been referred to adult services?

❑ YES

❑ NO

If YES, Fill in the "Transition" section; if NO go to page 5 (Section "Questions in case of failed referral")

#### **Transition**

1. Have there been informational meetings about the transition phase with the patient and family?

❑ YES

❑ NO

2. How old was the patient when preparation for transition to adult services began? ____________

3. How long did the transition phase take? (in months) ______________

4. How many meetings were conducted with the patient and his family?

________________________________

5. The timelines adopted for this patient were:

❑ Flexible (e.g., taking development into account)

❑ Based on strict criteria (e.g., age, severity of symptoms)

6. Was the time sufficient to allow a good transition from referral to exclusive patient care by the adult service?

❑ YES

❑ NO - how many more months would have been necessary? ________________

7. To which service was the patient referred? (more than one option can be selected)

❑ AMHS

- ❑ Territorial psychiatric service
- ❑ Private psychiatrist/psychologist/psychotherapist
- ❑ Attending physician
- ❑ Continued care in CAMHS
- ❑ I don't know
- ❑ Other Child Neuropsychiatry Service

8. Did you encounter any problems in sending it?
    - ❑ YES - specify: ____________________________________________________________
    - ❑ NO

9. Did you send information regarding the patient's medical history to the adult services to which they were referred?
    - ❑ YES
    - ❑ NO

If YES, specify how (more than one option can be selected):

- ❑ Information sheet/diagnostic report
- ❑ Phone
- ❑ Email
- ❑ Meeting between services
- ❑ Online meeting
- ❑ Other

10. With whom were the meetings conducted? (more than one option can be selected):
    - ❑ Interviews with the patient inherent to the pathway to be taken
    - ❑ Talks with the patient's family inherent to the path to be taken
    - ❑ Clinical case presentation and discussion between teams from the two involved services
    - ❑ Interview with adult service psychiatrist
    - ❑ Involvement of other professionals (curricular or support teachers, social services, educators)
    - ❑ None

11. Were these items present in the transition path? (more than one option can be selected)
    - ❑ Patient and family involvement
    - ❑ Transition planning, information sharing, and joint work between neuropsychiatrist and psychiatrist
    - ❑ Evaluation of procedures (e.g., updating diagnosis and drug treatment)
    - ❑ Continuity of care
    - ❑ Consideration of the most appropriate/specialized services in ADHD

12. In defining this patient's pathway, were there any special needs that led to the individualization of the pathway?

❑ YES

❑ NO

If YES, based on what (more than one option can be selected):

❑ Cognitive abilities and resources of the person

❑ Emotional and psychological state

❑ Needs related to long-term living conditions

❑ Social, economic, and family circumstances

❑ Responsibility of care

❑ Communication needs

❑ Support the person has available (including but not limited to family support)

❑ Educational and employment outcomes

❑ Community inclusion, emotional health, housing autonomy

13. Do you think the patient acquired new skills during the transition process?

❑ YES

❑ NO

If YES, which ones (more than one option can be selected):

❑ Knowledge of the disorder (onset, symptoms, persistence)

❑ Knowledge of comorbidities (possible other conditions concomitant to ADHD)

❑ Knowledge of primary treatments (psychological, educational, pharmacological)

❑ Knowledge of drugs (side effects, dosage)

❑ How to book appointments with the service (who to contact, what materials to bring)

❑ Development of independence and responsibility for care

14. Was the referral accepted immediately?

❑ YES

❑ NO - we had to fall back on - specify (e.g., private specialist): ____________________

❑ I don't know.

15. Did you ask for feedback from AMHS after referral?

❑ YES

❑ NO

16. Did you receive feedback from AMHS after referral?

- ❑ YES
- ❑ NO

17. Have any monitoring meetings been organized after the transition?

- ❑ YES - specify how many: ________________________________________
- ❑ NO

18. Following this patient's transition path, were there any particular obstacles or steps that could have been done differently?

- ❑ YES - specify:

  ______________________________________________________________________
- ❑ NO

#### Questions in case of failed referral

1. Did you attempt a referral?

- ❑ YES - to whom: ____________________________________________________________________
- ❑ NO

If YES - are you aware of why this submission was unsuccessful?

______________________________________________________________________

If NO - for what reason?

______________________________________________________________________

2. In following this patient's path after coming of age, were there any particular obstacles or steps that could have been taken differently?

- ❑ YES - specify:

  ______________________________________________________________________
- ❑ NO

#### 2.2 - Version for AMHS clinicians

*We kindly ask you to fill out this questionnaire for each ADHD patient who participated in the study and whom you followed up after transfer from the Child Neuropsychiatry Service (CAMHS). The person filling out the questionnaire must be the practitioner who has followed or is following the patient.*

The questionnaire is completed by (professional figure): ________________________________
Institution to which the compiler belongs: ________________________________
Compiler's email: ________________________________

Patient's date of birth: day |__|__|, month |__|__|, year |__|__|__|__|

Gender of the patient: M❑ F ❑

Patient diagnosis:

❑ ADHD - Predominant Inattention
❑ ADHD - Predominant hyperactivity/impulsivity
❑ ADHD - Combined

Indicate in which areas impairments were present when the patient was taken in care:

❑ Education/work
❑ Social/emotional relationships
❑ Impulsivity (e.g., money management, gambling, substance use, accidents/risk behaviors, sexuality)
❑ Emotionality (depression, anxiety, obsessions, fears/worries)
❑ Quality of life (sleep, nutrition, physical activity)

Indicate the patient's level of impairment at the time of admission:

❑ Mild
❑ Moderate
❑ Medium-severe
❑ Severe

Drug therapy at the time of admission:

❑ Yes - which one: ________________________________________________________________
❑ NO

#### TRANSITION QUESTIONS

1. Since you took in the patient, has his drug therapy been changed?

❑ YES – specify ____________________________________________________________

❑ NO

2. How long did you follow the patient with ADHD?

❑ Since his transfer (specify for how many months he followed the patient: ___________)

❑ I took him in later (specify for how many months he followed the patient: _________)

3. How old was the patient when they were taken in? ____________

4. Did you encounter any problems in receiving the patient?

❑ YES - specify: __________________________________________________________

❑ NO

5. Did you receive information regarding the patient's medical history from the CAMHS?

❑ YES

❑ NO

If YES, specify how (more than one option can be selected):

❑ Information sheet/diagnostic report

❑ Phone

❑ Email

❑ Meeting between services

❑ Online meeting

❑ Other

6. Were meetings conducted jointly with the CAMHS team before the transfer?

❑ YES

❑ NO

7. Were these items present in the transfer planning pathway? (more than one option can be selected):

❑ Patient and family involvement

❑ Joint planning of the timing of the transition, information sharing, and joint work between CAMHS and the patient's caregivers

❑ Evaluation of procedures (e.g., updating diagnosis and drug treatment)

❑ Consideration of specialized services in ADHD

❑ Involvement of other professionals (curricular or support teachers, social services, educators)

8. In defining this patient's pathway, were there any special requirements that led to its personalization?

- ❑ YES
- ❑ NO

If YES, according to (more than one option can be selected):

- ❑ Cognitive abilities and resources of the person
- ❑ Emotional and psychological state
- ❑ Needs related to long-term living conditions
- ❑ Social, economic, and family circumstances
- ❑ Responsibility of care
- ❑ Communication needs
- ❑ Support available to the person (including but not limited to family)
- ❑ Educational and employment outcomes
- ❑ Community inclusion, emotional health, housing autonomy

9. Do you think the patient acquired skills during the transition process?

- ❑ YES
- ❑ NO

If YES, which ones:

- ❑ Knowledge of the disorder (onset, symptoms, persistence)
- ❑ Knowledge of comorbidities (possible other conditions concomitant to ADHD)
- ❑ Knowledge of primary treatments (psychological, educational, pharmacological)
- ❑ Knowledge of drugs (side effects, dosage)
- ❑ How to book appointments with the service (who to contact, what materials to bring)
- ❑ Development of independence and responsibility for care

**FOLLOW-UP QUESTIONS**

10. Has the CAMHS requested feedback on the patient's condition?

- ❑ YES
- ❑ NO

11. Did you send feedback about this patient to the home CAMHS?

- ❑ YES
- ❑ NO

12. Were monitoring meetings held with the CAMHS after the transition process?

❑ YES - specify how many: _______________

❑ NO

13. were there any particular obstacles or steps that could have been taken differently in receiving this patient?

❑ YES - specify: ________________________________________________________________

❑ NO
