## Supplementary Material for "The Transition from adolescence to adulthood mental health services for young people with ADHD in Italy"

| ID | At age 18 services | Intermediate period other services | Current services | referred | outcome/reason for unsuccessful referral | impairment | need for support | employment | sentinel events | medications before transfer | current medications |
|---|---|---|---|---|---|---|---|---|---|---|---|
| 2 | AMHS | | AMHS | - | *already cared for by AMHS at 18* | moderate | no | unemployed | | drugs + therapy | no treatment |
| 3 | CAMHS + AMHS | CAMHS + AMHS | AMHS | - | *already cared for by AMHS at 18* | moderate | no | school | | drugs + therapy | no treatment |
| 4 | AMHS | | AMHS | - | *already cared for by AMHS at 18* | moderate | yes | school | | drugs + therapy | no treatment |
| 5 | AMHS | none | none | - | *already cared for by AMHS at 18* | moderate | no | school | | drugs + therapy | drugs + therapy |
| 7 | CAMHS | AMHS + private | AMHS | yes | *referral successful* | moderate | no | unemployed | | drugs + therapy | no treatment |
| 21 | CAMHS | AMHS + private | AMHS +private +center for drug addiction | no | lack of specialized service | moderate | yes | unemployed | | drugs + therapy | no treatment |
| 22 | CAMHS | private | CAMHS only for medications | no | deemed unnecessary | mild | no | school | | drugs + therapy | no treatment |
| 23 | CAMHS | AMHS | AMHS | yes | *referral successful* | medium/severe | no | unemployed | | drugs + therapy | no treatment |
| 8 | CAMHS | AMHS | AMHS | yes | *referral successful* | mild | yes | unemployed | | drugs + therapy | therapy |
| 11 | CAMHS | none | AMHS + private | yes | *referral successful* | mild | yes | work+school | yes | drugs | no treatment |
| 36 | CAMHS | CAMHS | territorial CAMHS | yes | *referral successful but not to AMHS* | moderate | yes | work | | drugs + therapy | therapy |
| 17 | CAMHS | none | none | yes | dropout, no compliance | moderate | no | work | | no treatment | therapy |
| 18 | CAMHS | none | none | no | lack of specialized service | mild | yes | work | | drugs + therapy | no treatment |
| 19 | CAMHS | none | none | no | lack of specialized service | moderate | yes | work | yes | drugs + therapy | therapy |
| 20 | CAMHS | none | none | no | lack of specialized service | moderate | no | work | | drugs + therapy | no treatment |
| 24 | CAMHS | none | none | no | dropout, no compliance | medium/severe | no | work | | drugs + therapy | no treatment |
| 9 | CAMHS | none | none | no | lack of specialized service | mild | yes | school | | drugs + therapy | no treatment |
| 10 | CAMHS | none | none | no | other CAMHS for complex clinical picture | moderate | no | work | | no treatment | drugs |
| 12 | CAMHS | none | none | yes | *referral successful* | mild | no | unemployed | | drugs | drugs + therapy |
| 13 | CAMHS | none | none | yes | *referral successful* | mild | yes | work+school | yes | drugs + therapy | drugs + therapy |
| 14 | CAMHS | none | none | no | lack of specialized service | moderate | yes | unemployed | yes | drugs + therapy | no treatment |
| 15 | CAMHS | none | none | no | lack of specialized service | moderate | no | work | yes | therapy | drugs + therapy |
| 29 | private | AMHS | private | no | dropout, no compliance | medium/severe | yes | work | | drugs + therapy | no treatment |
| 30 | private | AMHS + private | private | yes* | attempted but unsuccessful | medium/severe | no | school | | drugs + therapy | no treatment |
| 6 | private | none | none | yes* | attempted but unsuccessful | moderate | yes | school | | therapy | drugs |
| 31 | private | PRIVATE | private | yes* | attempted but unsuccessful | medium/severe | yes | work+school | | drugs + therapy | no treatment |
| 25 | private | none | none | no | patient chose a private professional | mild | no | work | | drugs + therapy | therapy |
| 1 | none | AMHS | AMHS | yes | attempted but unsuccessful | medium/severe | yes | school | | drugs + therapy | drugs |
| 16 | none | private | none | no | lack of specialized service | mild | yes | unemployed | | drugs + therapy | no treatment |
| 33 | none | none | none | no | deemed unnecessary | mild | no | work | yes | drugs + therapy | no treatment |
| 34 | none | none | none | no | deemed unnecessary | medium/severe | no | work | yes | drugs + therapy | no treatment |
| 35 | none | none | none | no | deemed unnecessary | mild | no | work | | drugs + therapy | no treatment |
| 32 | none | none | none | no | deemed unnecessary | mild | no | work | | drugs + therapy | no treatment |
| 26 | none | none | none | no | deemed unnecessary | mild | no | work | | drugs + therapy | no treatment |
| 27 | none | none | none | no | deemed unnecessary | mild | no | work | yes | drugs + therapy | no treatment |
| 28 | none | none | none | no | dropout, no compliance | mild | no | work | | drugs + therapy | no treatment |

Notes: "Intermediate period" refers to the time between young adults turning 18 and the year before the interview.

"Current" refers to the time of the interview and the year before the interview

* referral only attempted
